# Uncovering Hidden Pathogens in Culture Negative Cases: Enhancing Diagnostic Utility in Post-Neurosurgical Meningitis/Ventriculitis Using 16S Metagenomics

**DOI:** 10.64898/2025.12.05.25341694

**Authors:** Srinivasa Sundara Rajan Radhakrishnan, Veena Kumari Haradara Bahubali

## Abstract

Post-neurosurgical Meningitis/Ventriculitis remains a major clinical challenge, particularly in cases where conventional microbiological cultures yield negative results. Delayed or improper diagnosis can lead to sepsis or even death in patients. 16S metagenomics next-generation sequencing (mNGS) techniques have higher sensitivity and specificity than gold-standard techniques. This prospective study aimed to evaluate the ability of mNGS to detect pathogens in cerebrospinal fluid (CSF) in culture-negative cases. Forty-four CSF specimens collected from 44 patients with suspected central nervous system (CNS) infection were subjected to mNGS and microbial culture analysis in parallel. Among the 44 CSF specimens, 27 (79%) were NGS positive, which was higher than microbial culture 4 (9%). The sensitivity, specificity, positive predictive value (PPV), and negative predictive value (NPV) were 75%, 20%, 11%, and 85%, respectively. A high NPV aids in ruling out infection, and mNGS substantially improves pathogen detection in culture-negative cases. Further advances in sample processing methods are required to improve sensitivity and specificity.

**Importance:** This study aimed to evaluate the ability of mNGS to detect pathogens in the cerebrospinal fluid in cases of post-neurosurgical meningitis, which can have high mortality rates and severe sequelae. This prospective study of patients with suspected CNS infection found that mNGS has a higher sensitivity for detecting bacterial pathogens than the current gold standard microbial culture. It was the first time combining mNGS and microbial culture to verify the results using 16S mNGS. Although microbial culture was thought to be the gold standard for pathogen detection and diagnosis of infectious diseases, this study suggested that microbial culture of CSF is not the most appropriate way for diagnosing (CNS) infection. NGS should be recommended to be used in CSF for diagnosing CNS infection.

## Introduction

The gold standard microbial culture for identifying the causative agents of infectious diseases has not changed for many years, and it is time-consuming. This is also true for central nervous system (CNS) infections, such as meningitis, which have a low positive rate and cause permanent disabilities, such as brain damage, hearing loss, and learning disabilities. Despite dramatic advances in diagnostic technology, many patients suspected of having an infectious disease continue to receive empirical antibiotics instead of definitive antibiotics against the identified causative bacteria (1). Delayed diagnosis of bacterial infections can lead to sepsis and even death (2). Standard cultures have limitations regarding fastidious or even non-cultivable bacteria, and false-negative results can be obtained following antibiotic treatment.

CSF is considered a sterile fluid, and when infection is clinically suspected, the identification of microorganisms in CSF has traditionally relied on conventional bacterial cultures, which are tailored to identify specific human pathogens (3). Identifying a pathogen is vital, both in terms of guiding therapy and in characterizing prognosis. Due to the varying or non-specific symptoms of CNS infections in the early stages, it is difficult to make a definitive diagnosis, which leads to high mortality and morbidity. Inflammatory markers such as procalcitonin (PCT), interleukins, and total white cell count are used as indicators of infectious diseases but are not specific for CNS infections (4-7).

Therefore, accurate and efficient identification of pathogenic bacteria is essential for CNS infections. In recent years, metagenomic next-generation sequencing (mNGS) technology has been highly regarded for the diagnosis of pathogens in CNS infections (8), as it is a high-throughput sequencing technology that can be used for the unbiased detection of any pathogen without the need for sequence-specific amplification (9,10). mNGS has successfully detected a range of pathogens in patients with CNS infections. Several studies have shown that mNGS can detect pathogens in culture-negative samples, demonstrating its higher sensitivity than that of conventional methods.

This study aimed to perform 16s metagenomics sequencing on culture-negative CSF samples of patients with suspected bacterial meningitis using the MiSeq system, which offers a simpler NGS library preparation protocol, very low input DNA requirement, high-quality data, and faster turnaround time, and compare the clinical performance. We also used a simpler bioinformatics pipeline for bioinformatics analysis and developed a local shiny app for simpler and faster analysis.

## Material and methods

### Study population

In the present study, 44 patients with suspected CNS infections admitted to the Department of Neurosurgery, NIMHANS, from Nov 2023 to December 2024, were included in this prospective study. The inclusion criteria for the patients were as follows: (i) patients of all age groups who had undergone neurosurgery with typical symptoms (headache, fever, vomiting, altered sensorium, or limb weakness). The exclusion criteria were as follows: (i) patients with end-stage liver or renal disease or subdural or extradural hemorrhage and (ii) patients with incomplete clinical history. General clinical information (including age, gender, and results of laboratory tests) of all was obtained from the medical health records

### Ethics Statement

The research was performed in accordance with relevant guidelines.This study protocol was approved by the institutional ethics committee (IEC) (Basic Science & Neuroscience) of the National Institute of Mental Health & Neurosciences (NIMHANS). No. NIMHANS/IEC (BS & NS DIV)/2022 dated 26.05.2022

### Specimen collection and routine clinical laboratory tests

Leftover CSF from the specimens received in the laboratory for routine analysis was used for the study. The collected CSF was stored in a refrigerator at -80 °C for mNGS analysis. Routine investigations, including CSF biochemistry, CSF cytology, Gram staining, and CSF cultures, were performed for all patients.

### Nucleic acid extraction

DNA extraction was performed directly on the CSF samples without centrifugation. DNA was extracted using the QlAamp DNA kit (Qiagen, Germany) with slight modifications to the manufacturer’s instructions. Equal volumes of sample and lysis buffer were taken and incubated at 57 °C for 20 min following the addition of 20 µL proteinase K and 1 ml of chilled ethanol. Samples were incubated at room temperature for 1 min, centrifuged at 10,000 × g for 2 min, and standard wash procedures were followed. Finally, an optional step to enhance the total yield was performed, in which the elution buffer was heated to 60–70 and passed twice through the elution columns. All samples were eluted in 50 L Elution Buffer. DNA concentrations were determined using the Qubit Fluorometer with dsDNA HS Assay Kit (Thermo Fisher Scientific, USA)

### Microbial culture

Microbial culture was performed in the microbiological laboratory as a routine analysis. The CSF sample was inoculated into chocolate and blood agar plates and incubated at 35°C for 12–72 h. The final identification and characterization of pathogens was done by mass spectrometry (MS) (VITEK MS system, bioMerieux)

### Metagenomics next-generation sequencing

For 16s metagenomics next-genereation sequencing (m-NGS), DNA libraries were constructed using a 16S V4-V4 library preparation kit (Premas Life Sciences) according to the manufacturer’s instructions. All the constructed libraries were assessed for quality using a Qubit fluorometer (Thermo Fisher Scientific, USA) and an Agilent 2100 bioanalyzer (Agilent Technologies, CA). Samples were normalized to a concentration range of 2–4 nM, pooled, denatured with 0.2 N NaOH, and diluted with Illumina HT1 buffer to a 20 pM solution. A final loading volume of 600 l was loaded into an Illumina MiSeq cartridge and run overnight. To assess the quality control (QC) of the MiSeq run, we set a baseline acceptable Q30 (metric to assess base calling accuracy) score of 80% (i.e., 80% of each run’s calls had a 1 in 1,000 chance of being incorrect). DNA libraries were sequenced using the MiSeq Reagent Kit v3 (600-cycle) on the MiSeq platform (Illumina, CA).

### Development of a shiny app

For bioinformatics analysis, we developed a local, cross-platform graphical application in R using the Shiny framework to enable reproducible amplicon and metagenomic read processing without command-line requirements.

### Bioinformatics and statistical analysis

First, the output from Illumina was demultiplexed (converted from BCL to FASTQ) using the bcl2fastq software on a Linux-based system, which removed the dual index. To assess the quality of the fastq files, read quality control was performed using the FastQC online platform on the Galaxy server. Human reads were removed using Bowtie2 software. Sequencing data were analyzed, including filtering, error-model learning, ASV inference, chimera removal, and taxonomic assignment, using the denoising program DADA2 (11) and aligned to the SILVA 16S reference database (v. 138.2) (12, 13) to produce a 16S amplicon taxonomic table for downstream computational analysis. To remove environment and kit contamination, we used the decontam package in R (14, 15). Alpha diversity was evaluated using the Shannon, Simpson, and observed feature indices, and their significance was analyzed. Beta diversity was calculated using the Bray-Curtis method. All analyses were performed using R Studio version 4.3.

## Results

### Demographic and clinical data

Forty-four post-neurosurgical cases included in the study were numbered, and the details are shown in Table 1. Twenty-five males and 19 females, with a median age of 30 years, were suspected to have CNS infection but were culture-negative. The major clinical manifestations were headache and vomiting, and 4 had a history of head injury. Approximately 10% of patients did not survive treatment.

**Table 1:**
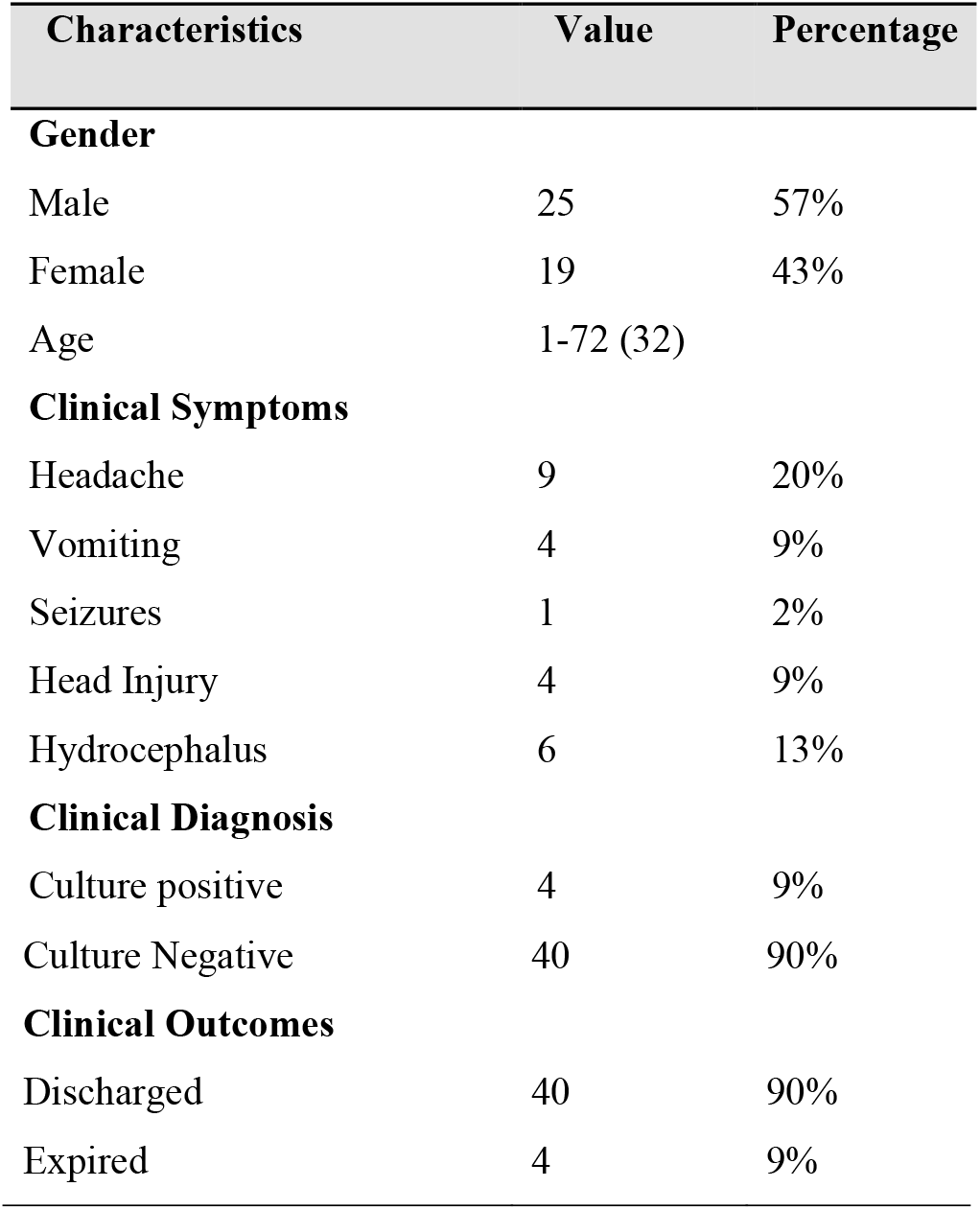
Demographic and clinical details.

| Characteristics | Value | Percentage |
| --- | --- | --- |
| Gender |  |  |
| Male | 25 | 57% |
| Female | 19 | 43% |
| Age | 1-72 (32) |  |
| Clinical Symptoms |  |  |
| Headache | 9 | 20% |
| Vomiting | 4 | 9% |
| Seizures | 1 | 2% |
| Head Injury | 4 | 9% |
| Hydrocephalus | 6 | 13% |
| Clinical Diagnosis |  |  |
| Culture positive | 4 | 9% |
| Culture Negative | 40 | 90% |
| **Clinical Outcomes** |  |  |
| Discharged | 40 | 90% |
| Expired | 4 | 9% |

### Clinical Laboratory Indicators

CSF samples were tested on the same day of collection for cell count and biochemistry and results are presented in the Table 2.

**Table 2:** Clinical laboratory indicators.

| Characteristics | Value (SD) |
| --- | --- |
| CSF Cell Count | 1251(± 1098.6) |
| Polymorphs | 900.7(±1168.4) |
| Lymphocytes | 97.21(±200.4) |
| Degenerated | 12 (±11.6) |
| **CSF Biochemistry** |  |
| CSF Glucose | 37.9(±29.3) |
| CSF Lactate | 79.8(±52.8) |
| CSF Protein | 487(±908.8) |
| Total Leukocyte Count | 13.1(±6.18) |

### Testing results of mNGS and culture

Among the 44 CSF specimens, 10 samples were not suitable for NGS analysis, of the 34 samples, 27 (79%) were NGS positive, and the detailed results are given in Table 3. The bacteria with the highest detection rates were *Acinetobacter baumannii* (4/34, 11.7%), followed by *Pseudomonas aeruginosa, Streptococcus parasuis, and uncultured Klebsiella sp. (*3/34, 8.8%). A total of 115 bacterial genera belonging to 15 phyla were detected using mNGS. One culture-positive case was undetected by mNGS. In a CSF specimen*, the culture results showed E. coli, whereas mNGS detected *Enterococcus faecalis* and *Streptococcus sp*. The sensitivity was found to be at 75%, and the specificity was 20%; the positive predictive value and negative predictive values were 85% and 11 %, respectively.

**Table 3:**
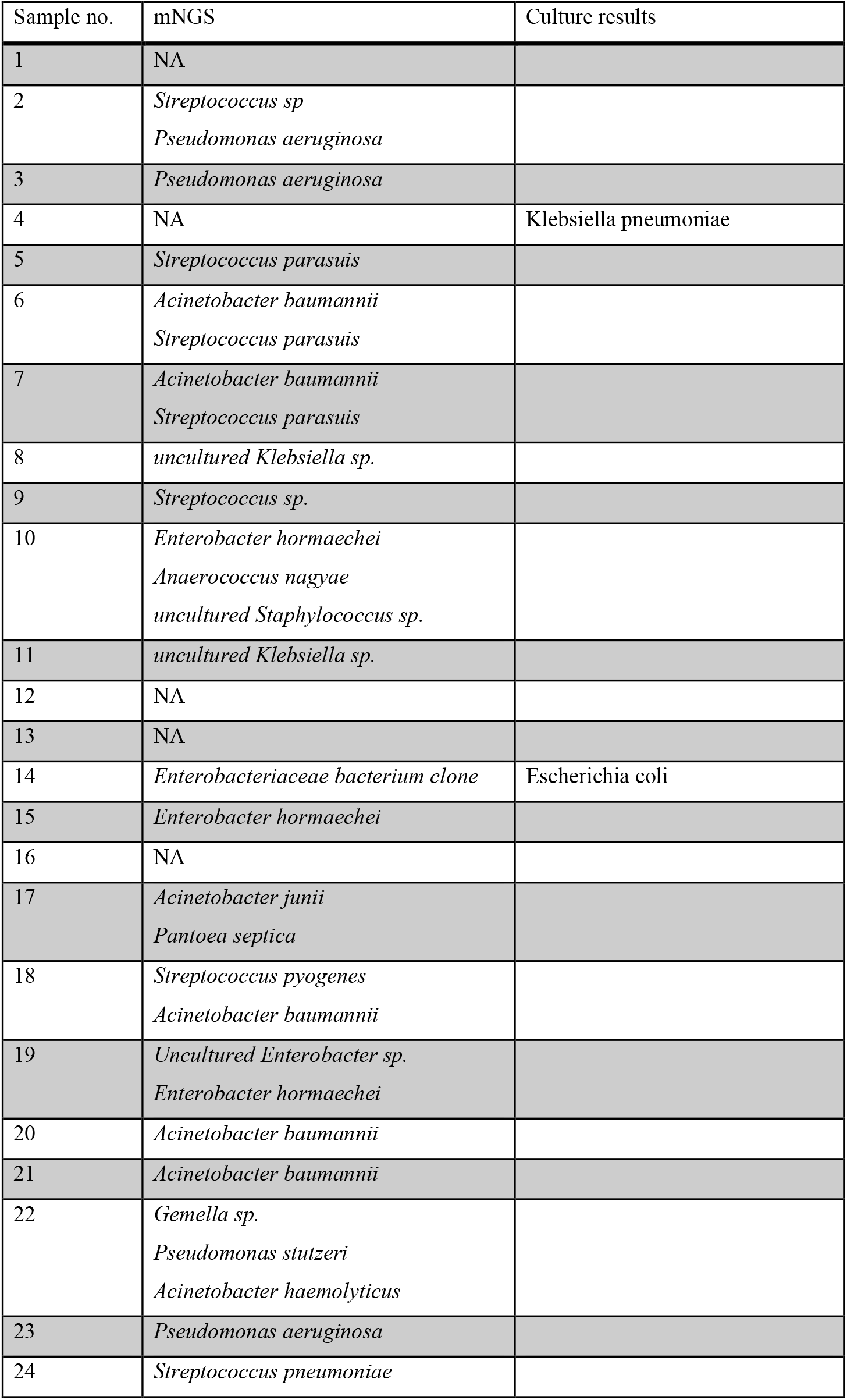
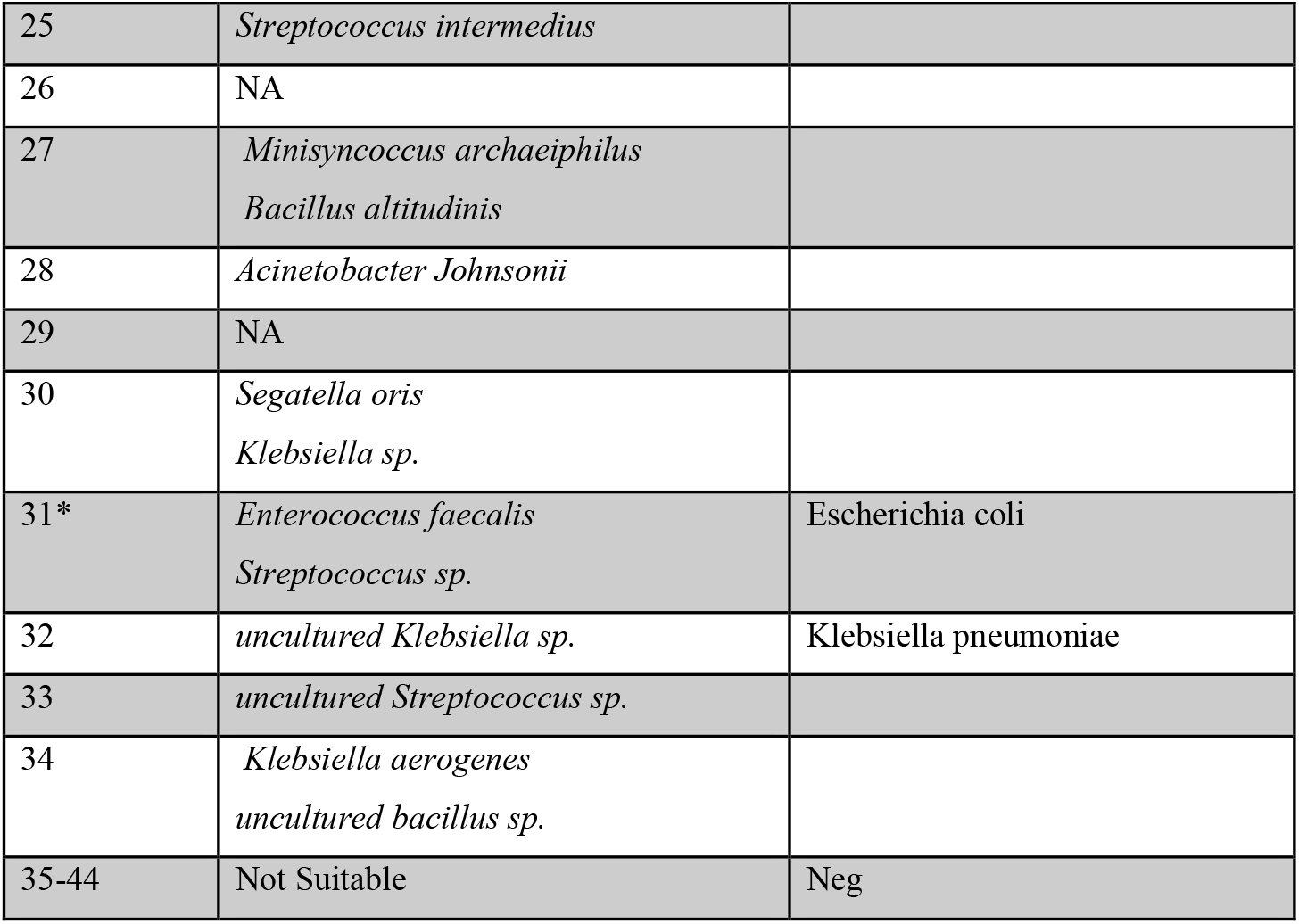
Results of mNGS and culture.

### Identification of bacteria

The workflow for bioinformatics analysis is shown in Figure 1. The sequences provided an overview of the pathogens in the CSF specimens. At the phylum level (Fig. 2), five known phyla were identified in the CSF specimens. *Pseudomonadota* and *Bacillota* were widely distributed and predominant among the pathogens, and at the genus level, the top genera with the highest abundance of sequences were *Acinetobacter* and *Streptococcus*.

**Fig 1:**
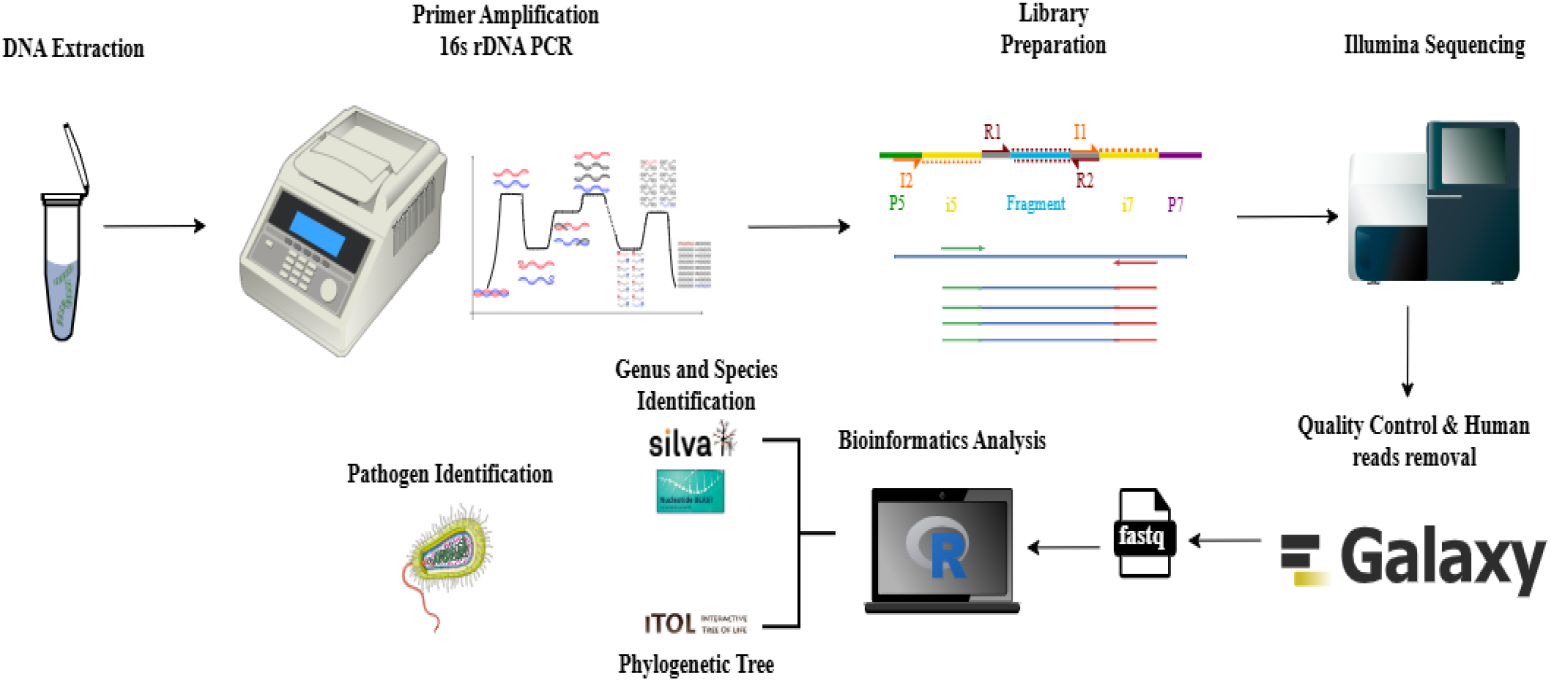
CSF workflow and bioinformatics pipeline for the diagnosis of meningitis

**Fig 2:**
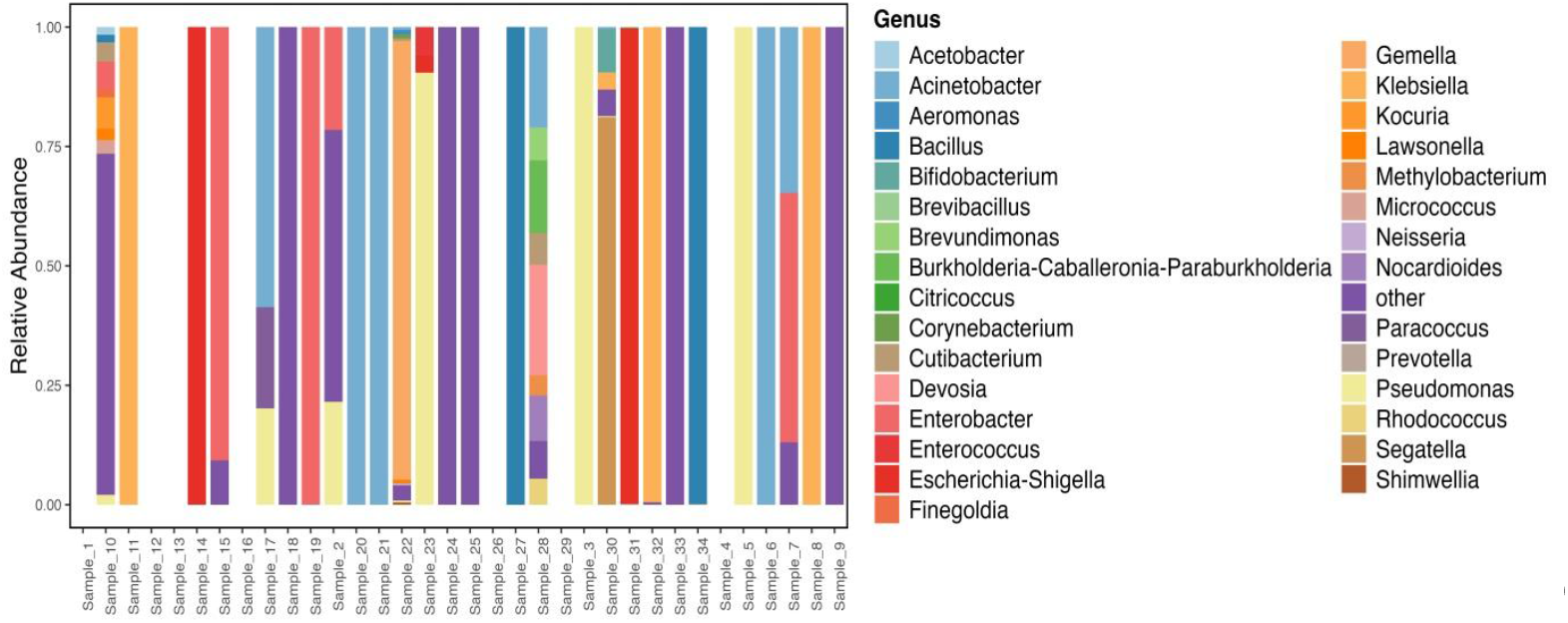
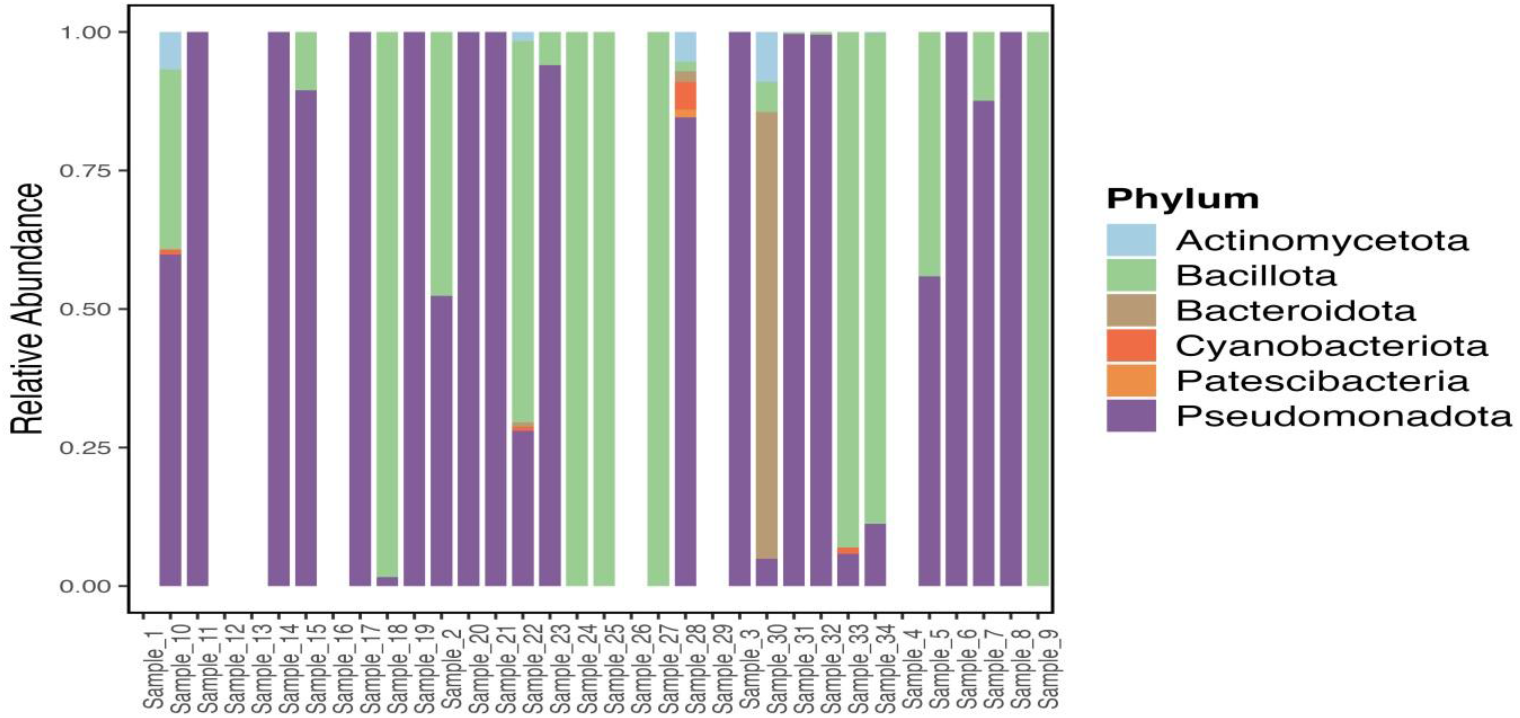
Genus and Phylum level microbial composition of cerebrospinal fluid (CSF) samples based on 16s rRNA sequencing. Relative abundance of the Top 30 genera is shown for each sample, with all remaining taxa grouped into the “other” category

### Phylogenetic tree analysis

Phylogenetic analysis showed that the bacterial taxa detected in the CSF were sparsely distributed across the tree without clustering into major enriched lineages (Fig. 3). The abundance bar/ring demonstrated that only a few genera contributed noticeably to the total microbial signal, with the majority of the detected genera present at extremely low relative abundances. This phylogenetic pattern indicates a weak or fragmented microbiome structure,

**Fig 3:**
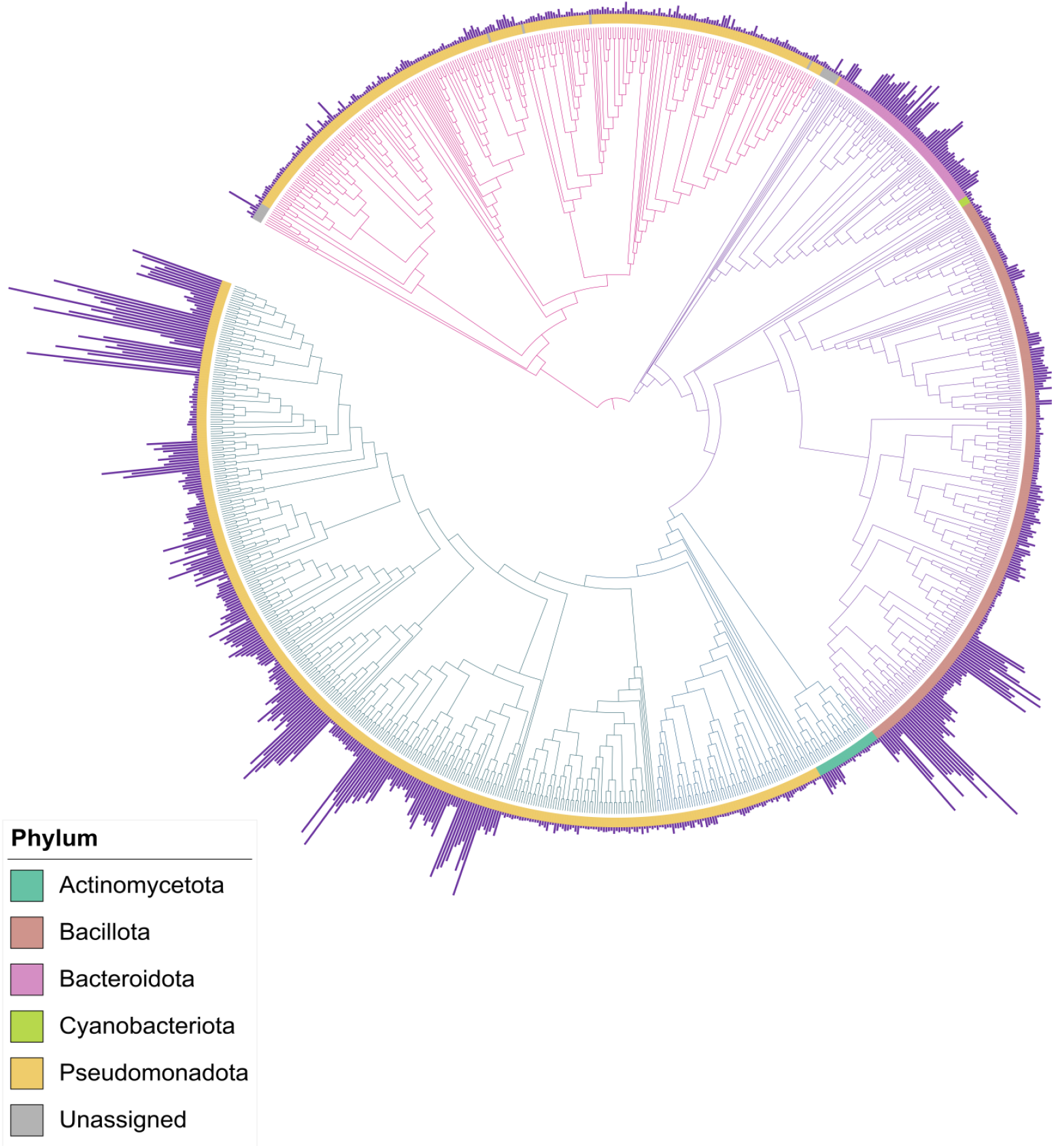
Phylogenetic tree based on Neighbour joining method using all 16s rRNA sequences. A midpoint-rooted phylogenetic tree was constructed using representative amplicon sequence variants (ASVs) aggregated at the genus level. The outer abundance bar/ring represents the cumulative relative abundance of each genus across all CSF samples.

which is expected in the CSF owing to its low microbial biomass. A small number of genera showed slightly higher abundance, but their placement on distant branches further supports the interpretation that no dominant phylogenetic clade was consistently enriched across the samples.

### Sample richness and diversity estimation

We performed a bioinformatics analysis of a large number of sequencing reads to evaluate the richness and diversity of CSF samples. We obtained a total of 1006 ASVs. Based on these data, we calculated the alpha and beta diversities. The plots (Fig. 4) show high variability and generally low diversity, which are common in CSF specimens. In the Shannon diversity plot, for most samples, the Shannon values were in the range of 0-2, indicating low or no diversity, and a few samples showed Shannon values in the range of 4, indicating a mixed bacterial signal, which is unusual in CSF. Simpson diversity, which focuses on evenness, showed values of approximately 0, indicating the dominance of one taxon. Few samples had mean abundances evenly distributed across taxa. The ASV for most of the samples ranges from 20-30, which is typical for CSF samples. The sequencing depth indicated that the samples were closely clustered with little beta separation. This analysis indicated that microbial variation was minimal.

**Fig 4:**
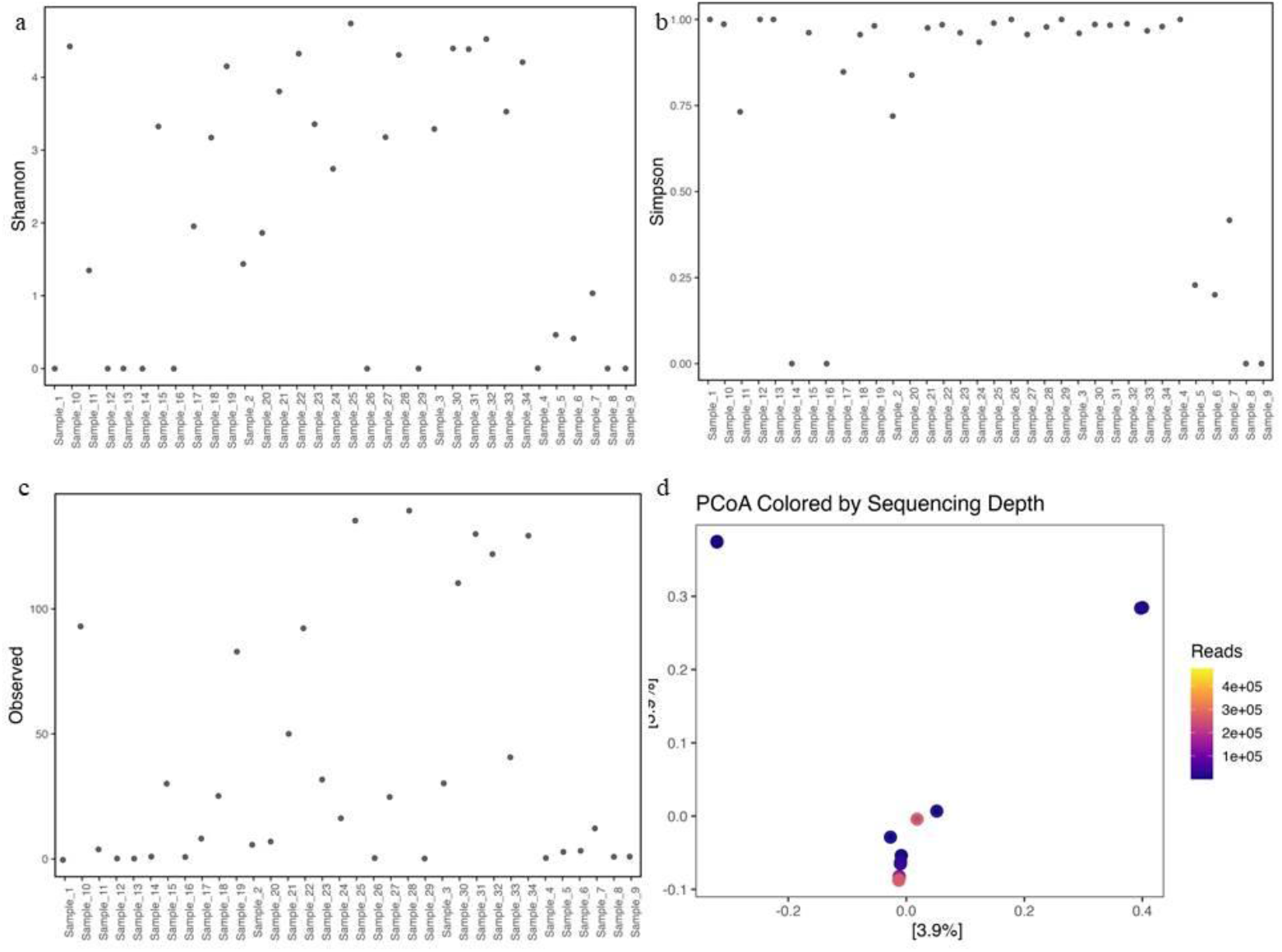
Alpha and beta diversity metrics of 16S rRNA sequencing data from cerebrospinal fluid (CSF) samples. a) Shannon diversity index b) Simpson diversity index c) Observed ASV richness d) Principal coordinates analysis (PCoA) based on Bray–Curtis distances, colored by sequencing depth

### Bioinformatics shiny app

We developed a user-friendly local Shiny app for bioinformatics analysis. The user must upload the *fastq* or *fastq*.*gz* to the interface and select whether it is a paired or single end, and set the parameters (truncLenF, truncLenR, maxEE, truncQ) manually, and the rest of the analysis is performed by the app. Reads will be filtered and trimmed based on user-defined parameters, followed by error model learning, and exact sequence variants (ASVs) will be inferred using *dada()* and merged for paired-end data. Chimeric sequences were removed using consensus-based detection methods. The resulting ASV table was exported with an ASV FASTA file. The taxonomic classification of ASVs was performed using *assignTaxonomy* with the SILVA reference database. () The application includes a validation module that verifies the integrity of the SILVA FASTA file by checking file accessibility, FASTA headers, file structure, and a live compatibility test to confirm the correct operation of *the assignTaxonomy function*. The results were returned as a taxonomy table containing kingdom to genus annotations (and species when references were included). For shotgun-style or non-denoised read classification, raw FASTQ files were processed using Kraken2 (16). Classification reports and label outputs were generated for each sample and summarized in tabular format. The application generates a structured output directory containing all intermediate and final files, including filtered FASTQs files, ASV tables, taxonomy tables, Kraken2 reports, and a downloadable ZIP archive.

## Discussion

CNS infections, such as meningitis, progress quickly and can be fatal. Accurate and prompt diagnosis is the premise of effective treatment. The traditional diagnostic paradigm often relies on positive culture and CSF smear results, which have low detection rates and can be delayed. Therefore, it is crucial to detect pathogens swiftly and effectively (9). Emerging mNGS offers advantages in terms of speed, detection rate, and identification of pathogens in unexplained infections (17).

Our prospective study is one of the few that combines mNGS and microbial culture and describes the microbiota present in CSF in cases of post-neurosurgical meningitis. We collected CSF samples from 44 patients suspected of having CNS infection who had undergone neurosurgeries and performed mNGS analysis on the samples. Of the 44 cases, 27 (79%) were NGS positive, and we had four (9%) culture-positive cases, of which one was found to be negative on mNGS. Acinetobacter baumannii (4/44, 11.7%) was the most common organism detected in the samples. The calculated sensitivity was 75%, which is high in the case of culture-negative cases, and the specificity was 20%, which is low because most patients would be on antibiotics, making it difficult to recover on culture. The positive and negative predictive values were 11% and 85%, respectively. The NPV was high, confirming the true negatives.

We identified S. parasuis, the second most common organism detected through mNGS, which is reported to cause meningitis, and the most common clinical syndrome of S. suis infection in humans is meningitis (18). One study demonstrated the capacity of S. parasuis strains to enter the CNS of infected mice (19). We found some rare bacteria in high abundance that can cause CNS infection, such as Cutibacterium acnes, and we found nine cases with C. acnes. It is mostly undiagnosed due to the difficulty in growing it in culture and is often reported as false-negative reports. It is reported to be the third most common cause of CSF shunt infection, alters the CSF proteome (20), and can form biofilms (21,22). We found two cases of Gemella sp. that can cause meningitis in patients with severe ENT conditions (23). Empedobacter brevis was identified in few samples, which is reported to be a rare cause of neonatal meningitis (24). Sagettella oris (Prevotella Oris) was observed in one case in low abundance, and it is reported to be a rare cause of brain abscesses. There were two case studies in which Prevotella oris was found to be of primary etiology, where it was unable to culture it, but confirmed after performing mNGS analysis (25, 26).

We performed alpha and beta diversity analyses to check the variability and diversity of the samples and found that they had high variability but low diversity, which is common in CSF because of low biomass.

While processing the specimen, we performed DNA extraction on neat CSF (without centrifugation) for NGS, which is reported to be the best method for pathogen detection (27). Of the 44 samples, 10 (22%) were not suitable for NGS analysis because the DNA concentration was low (>1ng/ul). Due to the characteristics of CSF samples, the concentration of nucleic acids may be very low, which impacts the ability to generate 16S rDNA amplicons, resulting in low-quality libraries and sequencing data (28). There were some contamination reads, which were mostly due to NGS reagents. It is reported and well documented that NGS reagents contain contaminating nucleic acids that interfere with the interpretation of results, especially among low-microbial-biomass CSF specimens (27). The identified taxa were predominantly bacteria that inhabit soil or water and are often linked to nitrogen fixation. This could be because nitrogen is commonly used as a substitute for air in ultrapure water storage tanks (29, 30).

Our study suggests that NGS sequencing can overcome the limitations of classical diagnostic testing methods by improving pathogen detection and has the advantage of detecting unknown or uncultivable bacterial species (31). Results from culture mostly depend on the number of viable bacteria, and the sensitivity of culture in bacterial meningitis is high, but it is diminished if CSF is obtained after initiation of antibiotics (32). mNGS may be used to evaluate the presence of many pathogens with a single test and has a high detection rate, even if only trace amounts of microbial DNA fragments are present (33).

We developed an intuitive Shiny application for processing NGS data using the DADA2 pipeline. The interactive interface allows users to upload files and execute the complete workflow without command-line steps. To the best of our knowledge, this is the first locally run, privacy-preserving, and user-friendly graphical tool for DADA2-based NGS data analyses.

However, our study has some disadvantages: (i) due to its targeted nucleic acid amplification of bacteria, it could not be used for identification of viral CNS infection; (ii) the volume of CSF available for analysis was relatively low (200 µL), which could hinder multiple DNA extraction for high-quality DNA; (iii) 16s mNGS could not generate data about clinical epidemiological information including strain typing, virulence, and drug-resistance genes.

## Data Availability

All data produced in the present study are available upon reasonable request to the authors

## Conclusion

In this study of post-neurosurgical meningitis, we demonstrated that compared to traditional microbiological diagnostic methods, mNGS substantially enhances pathogen detection and identifies novel and uncultivable pathogens in culture-negative CSF specimens. mNGS identified pathogens in 75% of clinically suspected CNS infections. Integrating mNGS with conventional microbiology and clinical judgement can significantly improve diagnostic accuracy and guide optimal management. Our findings highlight the potential of mNGS to overcome major limitations of culture, particularly in patients pre-treated with antibiotics, and to uncover pathogens that may otherwise remain undetected in culture.

## Authors Contribution

Conceptualization: Veena Kumari HB; Data Curation: Srinivasa Sundara Rajan R; Methodology: Veena Kumari HB, Srinivasa Sundara Rajan R; Software: Srinivasa Sundara Rajan R; Formal analysis and investigation: Veena Kumari HB; Writing - original draft preparation: Srinivasa Sundara Rajan R; Writing - review and editing: Srinivasa Sundara Rajan R, Veena Kumar HB,

## Statements and Declarations

The authors declare that they have no known competing financial interests or personal relationships that could influence the work reported in this study

